# Analysis of associations between polygenic risk score and COVID-19 severity in a Russian population using low-pass genome sequencing

**DOI:** 10.1101/2023.11.20.23298335

**Authors:** Arina V. Nostaeva, Valentin S. Shimansky, Svetlana V. Apalko, Ivan A. Kuznetsov, Natalya N. Sushentseva, Oleg S. Popov, Anna Y. Anisenkova, Sergey V. Mosenko, Lennart C. Karssen, Yurii S. Aulchenko, Sergey G. Shcherbak

## Abstract

The course of COVID-19 is characterized by wide variability, with genetics playing a contributing role. Through large-scale genetic association studies, a significant link between genetic variants and disease severity was established. However, individual genetic variants identified thus far have shown modest effects, indicating a polygenic nature of this trait. To address this, a polygenic risk score (PRS) can be employed to aggregate the effects of multiple single nucleotide polymorphisms (SNPs) into a single number, allowing practical application to individuals within a population. In this work, we investigated the performance of a PRS model in the context of COVID-19 severity in 1,085 Russian participants using low-coverage NGS sequencing. By developing a genome-wide PRS model based on summary statistics from the COVID-19 Host Genetics Initiative consortium, we demonstrated that the PRS, which incorporates information from over a million common genetic variants, can effectively identify individuals at significantly higher risk for severe COVID-19. The findings revealed that individuals in the top 10% of the PRS distribution had a markedly elevated risk of severe COVID-19, with an odds ratio (OR) of 2.1 (95% confidence interval (CI): 1.4–3.2, p-value = 0.00046). Furthermore, incorporating the PRS into the prediction model significantly improved its accuracy compared to a model that solely relied on demographic information (p-value < 0.0001). This study highlights the potential of PRS as a valuable tool for identifying individuals at increased risk of severe COVID-19 based on their genetic profile.

## INTRODUCTION

COVID-19, also known as coronavirus infection, is a contagious illness caused by the severe acute respiratory syndrome-coronavirus-2 (SARS-CoV-2). The majority of individuals who contract the virus exhibit mild to moderate respiratory symptoms and can recover without requiring specific medical treatment. However, in certain cases, the disease can manifest in a severe form, requiring medical intervention [1,2].

Apart from external factors like virus characteristics and the effectiveness of public health, certain host-related factors such as older age, male gender, and pre-existing chronic diseases like hypertension and diabetes have been associated with susceptibility and severity of COVID-19 [3,4]. However, these risk factors alone cannot fully explain the wide variation observed in the disease severity. The course of COVID-19 can range from asymptomatic cases to acute respiratory distress and even death [5,6]. Early in the pandemic, it was noted that clinical factors alone were insufficient to account for the variability in disease severity across individuals, as severe cases were observed in young people without apparent predisposing factors, often within families [7]. This suggests that human genetics may play a role in the development of the disease.

To gain insights into the aetiology of COVID-19, large-scale genetic association studies incorporating both rare and common genetic variants have employed various study designs. These investigations, along with subsequent follow-up studies, have expanded our understanding of the disease and provided potential avenues for its treatment. The COVID-19 Host Genetics Initiative (HGI) was established to identify genetic loci that impact the severity and susceptibility of COVID-19 [8]. This global effort aims to conduct a meta-analysis of multiple COVID-19 genome-wide association studies (GWAS), and to identify significant single nucleotide polymorphisms (SNPs) associated with infection, hospitalization, and mortality. Through comparisons of genomes of millions of COVID-19 patients and healthy individuals, these studies have implicated genetic variants in 13 loci associated with the severity of the disease [9]. The COVID-19-associated genetic variants could be related to the regulation of processes such as innate antiviral defence signalling, regulation of inflammatory organ damage, and upregulation of cell receptors [10]. Modulation of these pathways can impact susceptibility to infection and subsequent disease manifestation [11].

The effects of individual genetic variants identified so far are generally small, consistent with the polygenic architecture of this trait. An individual who tests negative for a specific risk variant may still have a high genetic risk due to other unmeasured genetic factors. While each single variant only explains a small portion of the risk for severe COVID-19, combining multiple genetic variants into a polygenic risk score (PRS) can offer a better prediction of the risk. PRS allows for the aggregation of the effects of multiple SNPs into a single score, which can be practically applied to individuals within a population [12]. Conventionally, a polygenic score is defined as a weighted linear combination of allele counts for SNPs observed in an individual’s genome. The PRS model consists of the weights of a set of SNPs, with the weights proportional to the estimated effects of the SNPs on the trait being studied [13].

Modern polygenic risk score models for human traits are typically estimated using summary statistics obtained from a genome-wide association meta-analysis (GWAMA) and a reference panel reflecting linkage disequilibrium (LD) in the population [13,14]. Over the past decade, PRS predictive performance has significantly improved due to larger GWAS sample sizes and advancements in methods for variable selection and effect estimation [15–24]. Polygenic scores can be utilized to rank individuals within a group based on their genetic predisposition to a disease [25–27]. This approach considers an individual’s genetic predisposition relative to the genetic predisposition of others in the same group, often expressed as a percentile representing where the individual’s PRS falls within the overall distribution of the group’s PRS.

Several studies have explored the development and application of PRS using variants associated with COVID-19, revealing clear associations between PRS and the risk of severe disease. However, most PRS models have been applied to cohorts consisting predominantly of individuals of Western European ancestry [28–31]. Using 1,582 SARS-CoV-2 positive participants from the UK Biobank (1,018 with severe COVID-19 and 564 without severe COVID-19) and 64 SNPs for PRS calculation, Dite et al. developed and validated a clinical and genetic model for predicting the risk of severe COVID-19. Only 13% of participants from this study were non-white, and PRS alone had an area under the receiver operating characteristic curve (AUC) of 68% [31].

While one recent study included African and South Asian groups, the associations with COVID-19 outcomes were limited by applying a PRS based on only six SNPs [32]. Another study that considered non-Western European populations was constrained by its focus on a specific Russian cohort (athletes) and also included only six genetic polymorphisms in the PRS assessment [33]. The multi-ethnic approach implemented in a very recent paper using UK biobank data, allowed the applicability of PRS, based on 17 SNPs, to diverse populations, with the severity model performing well within Black and Asian cohorts [34,35]. Overall, results highlight the potential of PRS as a predictive marker for disease severity and provide further support for its application in risk stratification and personalized healthcare approaches in the context of COVID-19.

Our study aimed to investigate the performance of the PRS model in the Russian population. The genomes of study participants (336 individuals with severe COVID-19 and 704 with moderate or without disease) were assessed using low-coverage (with mean depth x3) sequencing. Next, we developed a genome-wide PRS model for COVID-19 severity using the summary statistics from the COVID-19 host genetics initiative consortium. We demonstrated that PRS, incorporating information from more than a million common genetic variants, for COVID-19 severity can identify individuals with markedly elevated risk of severe COVID-19 course: OR = 2.1 (95% confidence interval (CI): 1.4–3.2, p-value = 0.00046) for individuals in the top 10% of the PRS distribution, and produces a significant improvement in the quality of prediction (p-value < 0.0001) compared to a model including only demographic information.

## RESULTS

### Participant Characteristics

The participants of the study were the patients of the infectious disease department of the St. Petersburg State Health Care Institution “City Hospital No. 40, Kurortny District” who were admitted for treatment with coronavirus infection (confirmed by polymerase chain reaction), and healthy individuals. Healthy individuals are defined as people who did not require COVID-19 medical treatment at the time of the study (between April 2020 and March 2022).

Table 1 shows the participants’ characteristics. Of the 1040 participants, 458 (44%) were female, with a mean age of 60 years, while for 582 (56%) males the mean age was equal to 56 years. Overall, 859 (82%) of all participants had COVID-19, of which 336 (39% from participants with COVID-19) had severe COVID-19. Separation according to the severity of the disease was carried out according to the following criteria: the case group included 336 patients (208 men and 128 women, 63±15 years) with lung damage more than 50% (computed tomography (CT)-3 and CT-4), the control group included 704 patients (374 men and 330 women, 56±16 years), with lung damage less than 50% or without COVID-19.

**Table 1.** The demographic and clinical characteristics of the participants.

| Characteristics | Male | Female |
| --- | --- | --- |
| Mean age (sd) | 56 (15) | 60 (16) |
| Healthy individuals | 110 | 71 |
| Patients required treatment by severity | CT-1: 77<br>CT-2: 187<br>CT-3: 207<br>CT-4: 1 | CT-1: 67<br>CT-2: 192<br>CT-3: 125<br>CT-4: 3 |
| Outcome | death: 90<br>recovery or no disease: 492 | death: 50<br>recovery or no disease: 408 |

Abbreviations: *CT* computed tomography, where CT-1 – mild form of pneumonia with areas of “frosted glass”, the severity of pathological changes less than 25%; CT-2 – moderate pneumonia, 25–50% of lungs are affected; CT-3 – moderately severe pneumonia, 50–75% of lungs are affected; CT-4 – severe form of pneumonia, >75% of lungs are affected.

### Low coverage sequencing and imputation

For all samples low coverage sequencing, also called LP-WGS (low-pass whole genome sequencing), was performed with a depth of 3x genome coverage. LP-WGS is the type of WGS with genome coverage from 0.5x to 5x [36,37]. Due to low-coverage data often having poor genotype quality and resulting in high missing genotype rates, the genotype likelihoods (GL) need to be updated using a reference panel for more accurate genotype imputation [38,39]. We used a recent method called GLIMPSE, which performs haplotype phasing and genotype imputation for LP-WGS data through a Gibbs sampling procedure, leading to improved accuracy [38]. As a reference panel, we used the 1000 Genomes data [40]. To evaluate the efficiency of LP-WGS within PRS, we calculated PRS values for a sample (not included in the study population) sequenced 41 times (in each of the batches to control the quality of the sequencing process). The coefficient of variation (CV) for PRS values was equal to 0.5% demonstrating a good method performance.

### Overview of the approach

Computation of PRSs requires both genotype data of target individuals and the PRS model. To build the PRS model we used summary statistics from the COVID-19 Host Genetics Initiative consortium (release 7) [8]. These results were obtained by the meta-analysis, which combined the results of 60 individual studies from 25 countries, with a total of 18,000 severe cases of COVID-19 and more than a million controls who either did not have a severe disease course or were not affected by COVID-19 during the study period. From the obtained summary statistics, we generated the PRS model using the Bayesian approach SBayesR with default parameters, implemented in the GCTB software [19,41,42]. Finally, we calculated individual PRS values using the PRS model (Fig. 1, Methods).

**Figure 1.**
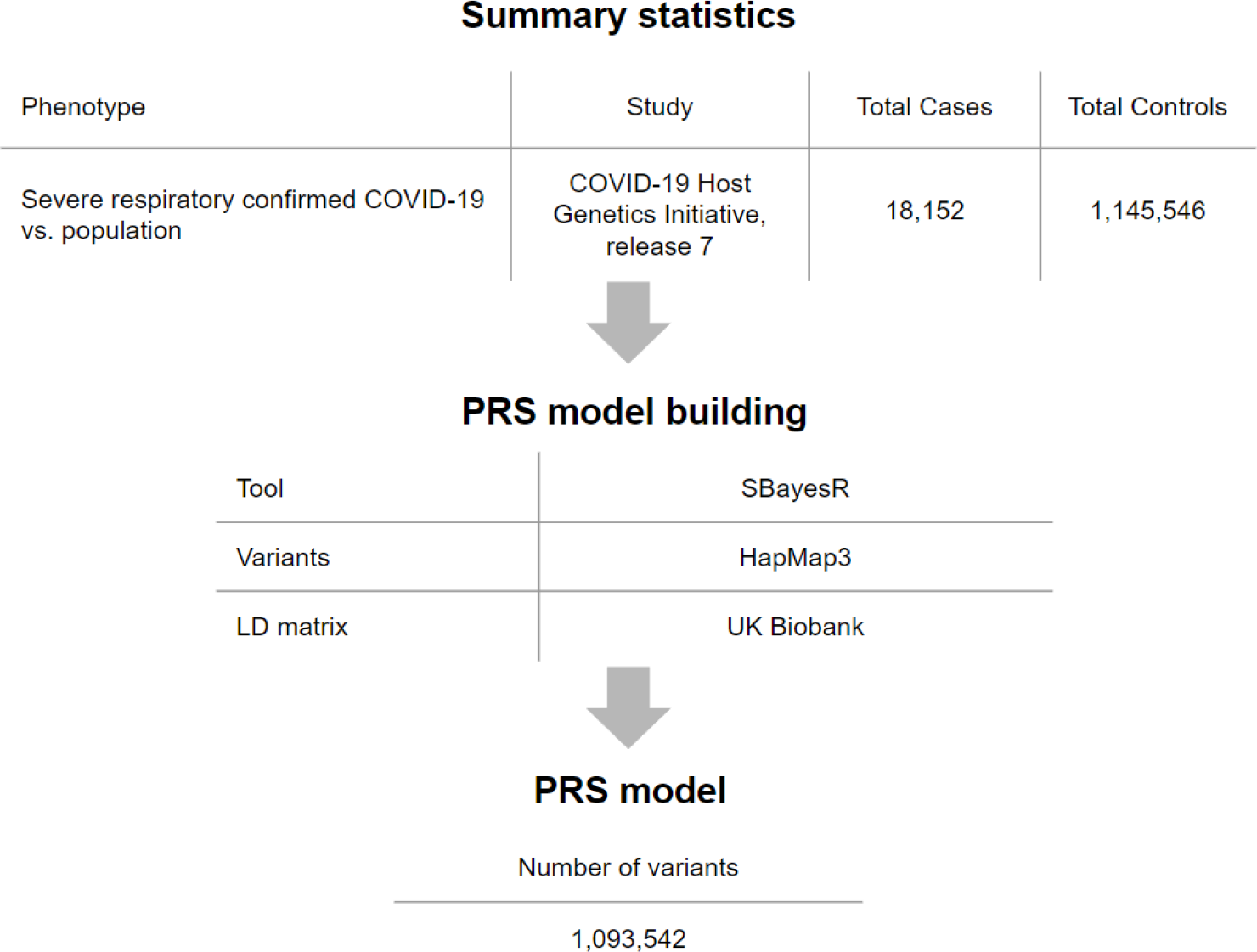
Study design and workflow. The PRS model for COVID-19 severity was derived by combining summary association statistics from the COVID-19 Host Genetics Initiative consortium and a linkage disequilibrium reference panel of 50,000 individuals of European ancestry from the UK Biobank data set. As a computational algorithm, SBayesR was used, which is a Bayesian approach to calculate a posterior mean effect for all variants based on a prior (effect size in the previous GWAS) and subsequent shrinkage based on linkage disequilibrium. The PRS model was restricted by the list of variants from HapMap3 and included about one million variants.

### Testing associations between PRS and severe COVID-19

We compared the distributions of PRS values between severe cases and the control group combining the milder forms of COVID-19 and healthy individuals (Fig. 2). Comparison of the mean PRS values, performed using Student’s t-test for two independent samples, showed significant difference (p-value = 1.8e-07).

**Figure 2.**
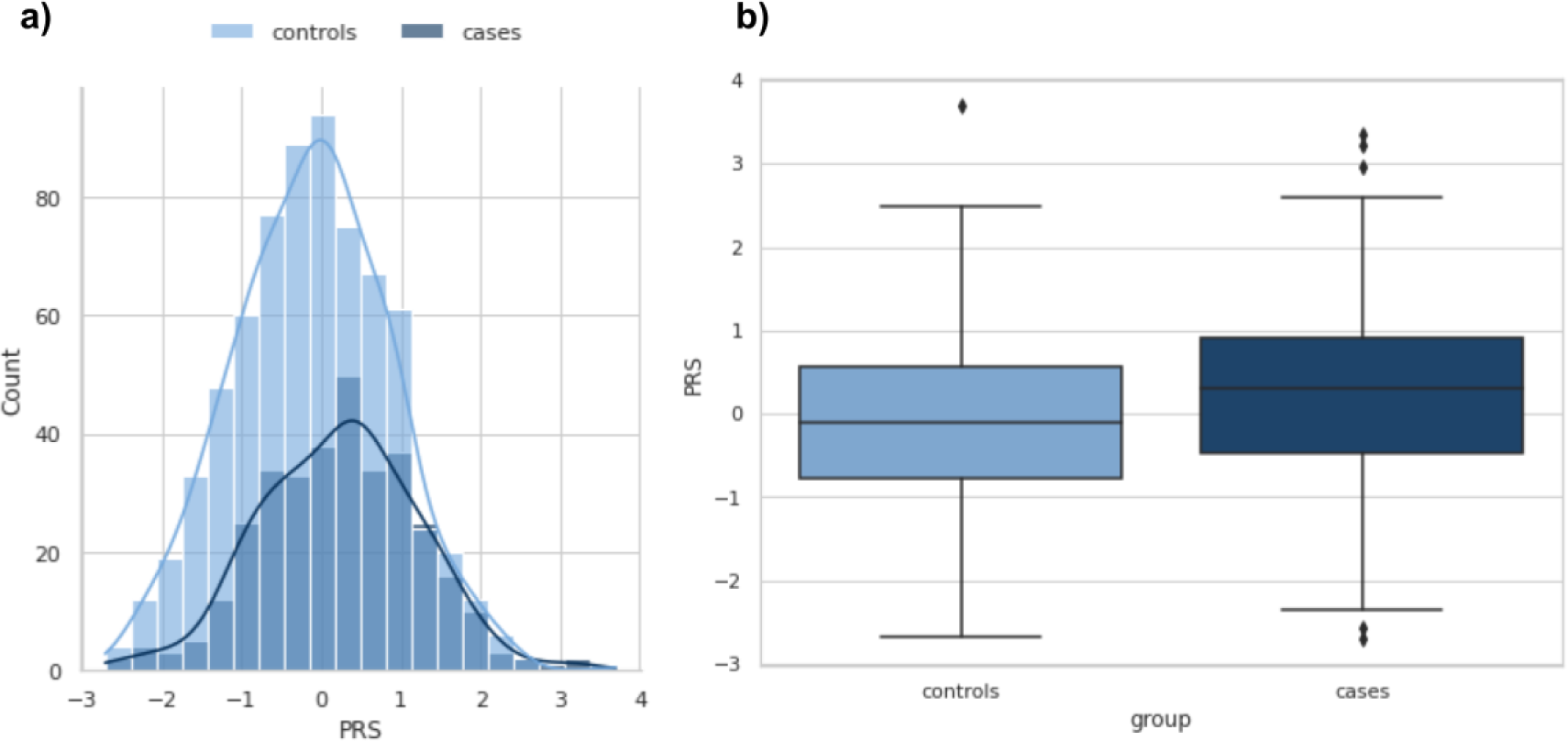
Comparison of distributions of PRS values between the groups with and without severe COVID-19. a) Distribution of PRS in the groups with (N_cases_ = 336) and without (N_controls_ = 704) severe COVID-19. The x-axis represents PRS, with values scaled to a mean of 0 and a standard deviation of 1 (in the total sample) to facilitate interpretation. b) PRS values among cases versus controls. Within each box plot, the horizontal lines reflect the median, the top, and bottom of each box reflect the interquartile range, and the whiskers reflect the rest of the distribution, except for points that are determined to be “outliers” .

Across the study population, the PRS was normally distributed with the risk of severe COVID-19 rising in the right tail of the distribution, from 15% in the lowest decile to around 50% in the highest decile (Fig. 3).

**Figure 3.**
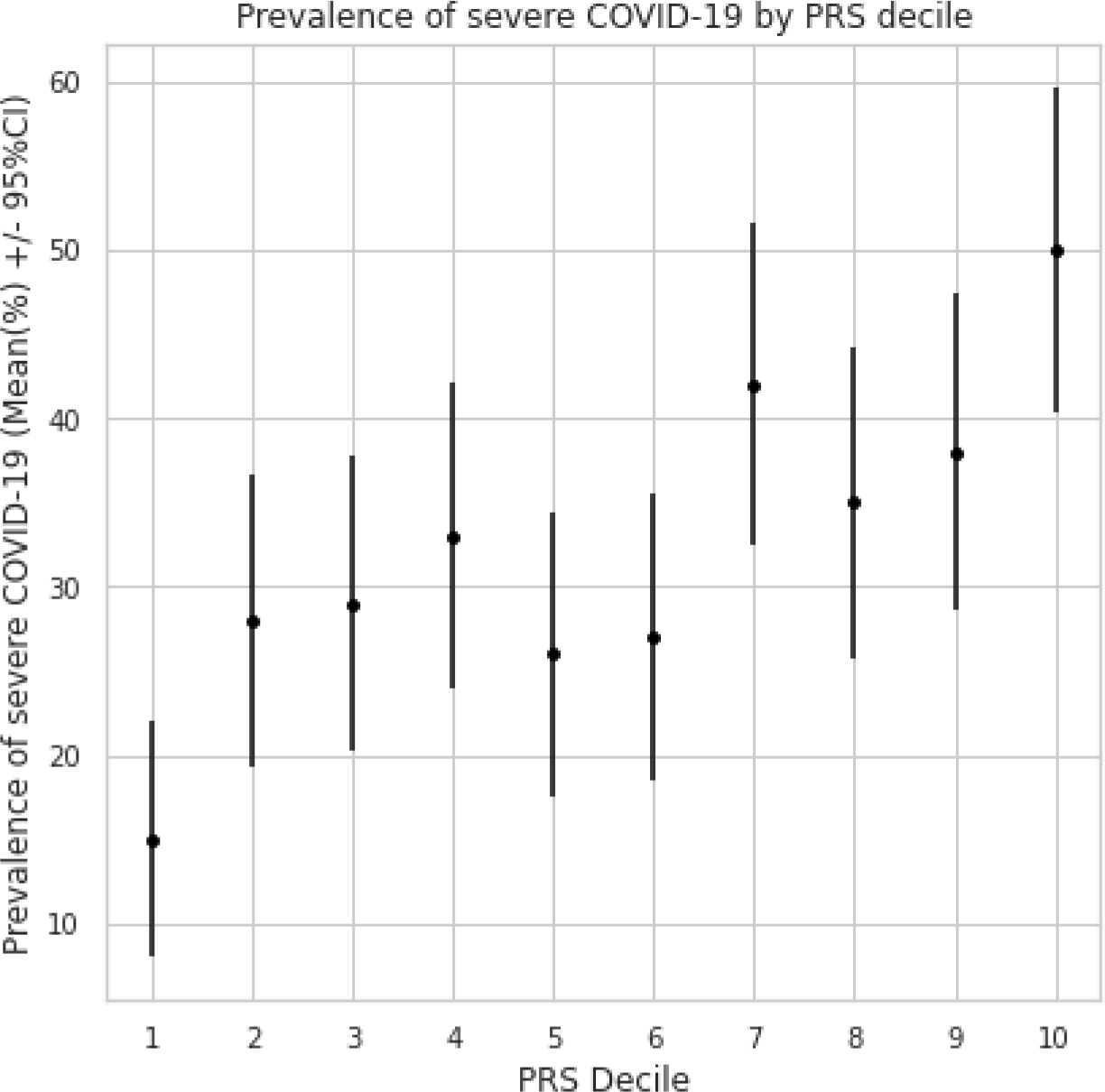
Prevalence of the severe COVID-19 according to PRS decile. All participants (N = 1,040) were stratified by decile of the PRS distribution. The average prevalence in percent and 95% CI within each decile are displayed.

Next, we found that 20% of the population with the highest PRS values had inherited a genetic predisposition that conferred OR = 1.66 for severe COVID-19 (95% CI: 1.2–2.3, p-value = 0.0013) in comparison with the whole cohort. The 10% of the population with the highest PRS values had an OR = 2.1 for COVID-19 (95% CI: 1.4–3.2, p-value = 0.00046).

### Evaluating the relationship between PRS and COVID-19 outcome

The severe form of the disease is associated with an increased risk of death. To assess how much the risk of death is associated with an increased PRS value, we calculated the odds ratio (OR) for death between the group with the highest PRS values (10%) and all others. The resulting OR was 1.8 (95% CI: 1.05–3.04) with p-value = 0.026. Thus, in the group with the highest PRS values, the probability of death due to severe disease was almost doubled.

We also compared the mean PRS values for groups with different COVID-19 outcomes (death vs no death or no disease). Results showed significant difference in mean PRS (p-value = 0.005, Fig. 4).

**Figure 4.**
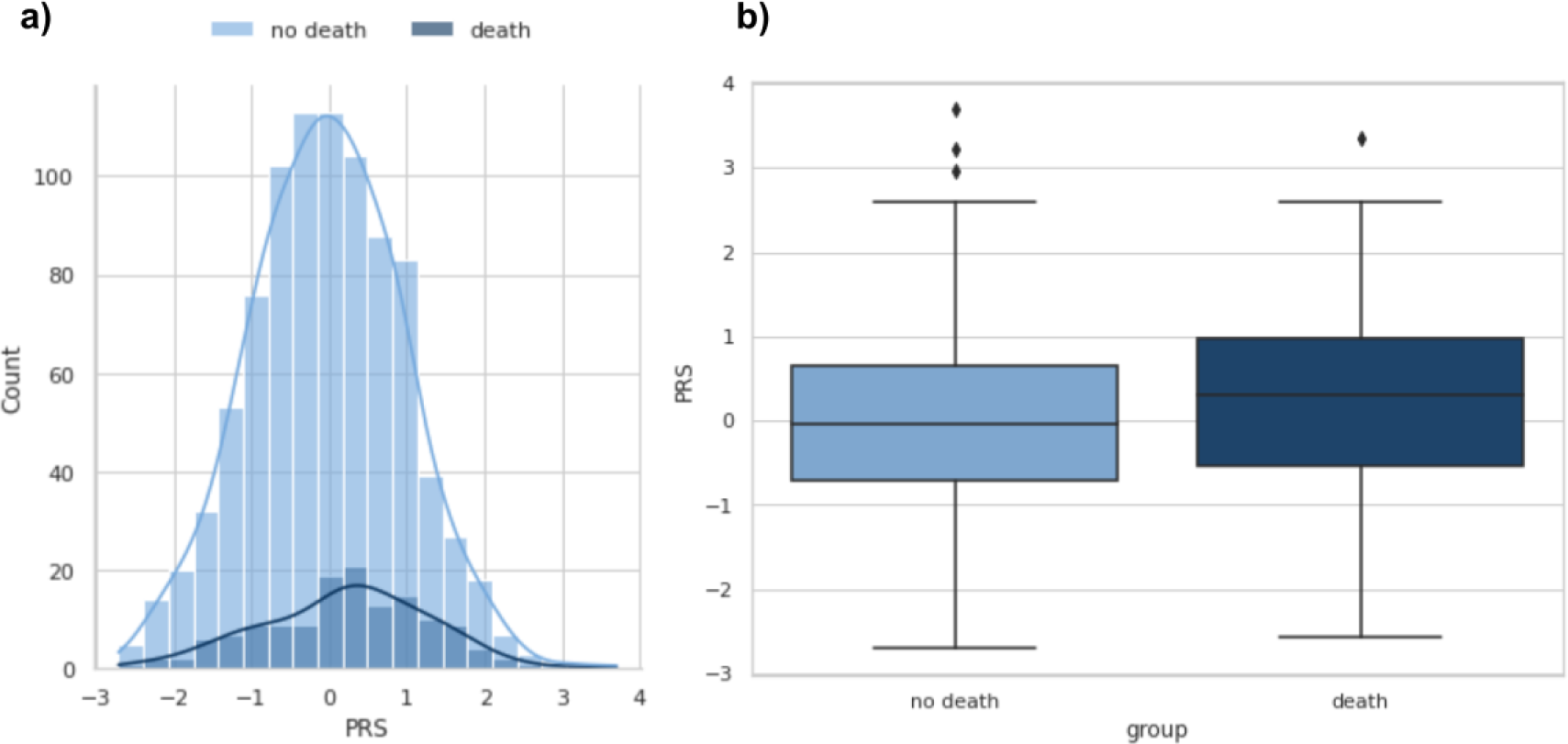
Comparison of distributions of PRS values between the groups with and without death outcome. a) Distribution of PRS in the groups with (N_death_ = 140) and without (N_no death_ = 900) death outcome of COVID-19. The x-axis represents PRS, with values scaled to a mean of 0 and a standard deviation of 1 (in the total sample) to facilitate interpretation. b) PRS values among cases versus controls. Within each box plot, the horizontal lines reflect the median, the top, and bottom of each box reflect the interquartile range, and the whiskers reflect the rest of the distribution, except for points that are determined to be “outliers” .

Next, we hypothesized that PRS for severe COVID-19 would be associated with a higher risk of severe COVID-19 in early age. In Kaplan–Meier analyses, which is a non-parametric statistic used to estimate the survival function from lifetime data, we divided the sample into three groups: 10% of all individuals with the highest PRS values, 10% of all individuals with the lowest PRS values and the rest (Fig. 5). The analysis showed that people from the group of high PRS values start to have increased risk in comparison with other groups already before the age of 40 years (p-value < 4.9e-9 for the log rank test). For example, the average risk of a severe course, which is reached at the age of 60 years, in the group with the highest PRS is reached before 50 years of age.

**Figure 5.**
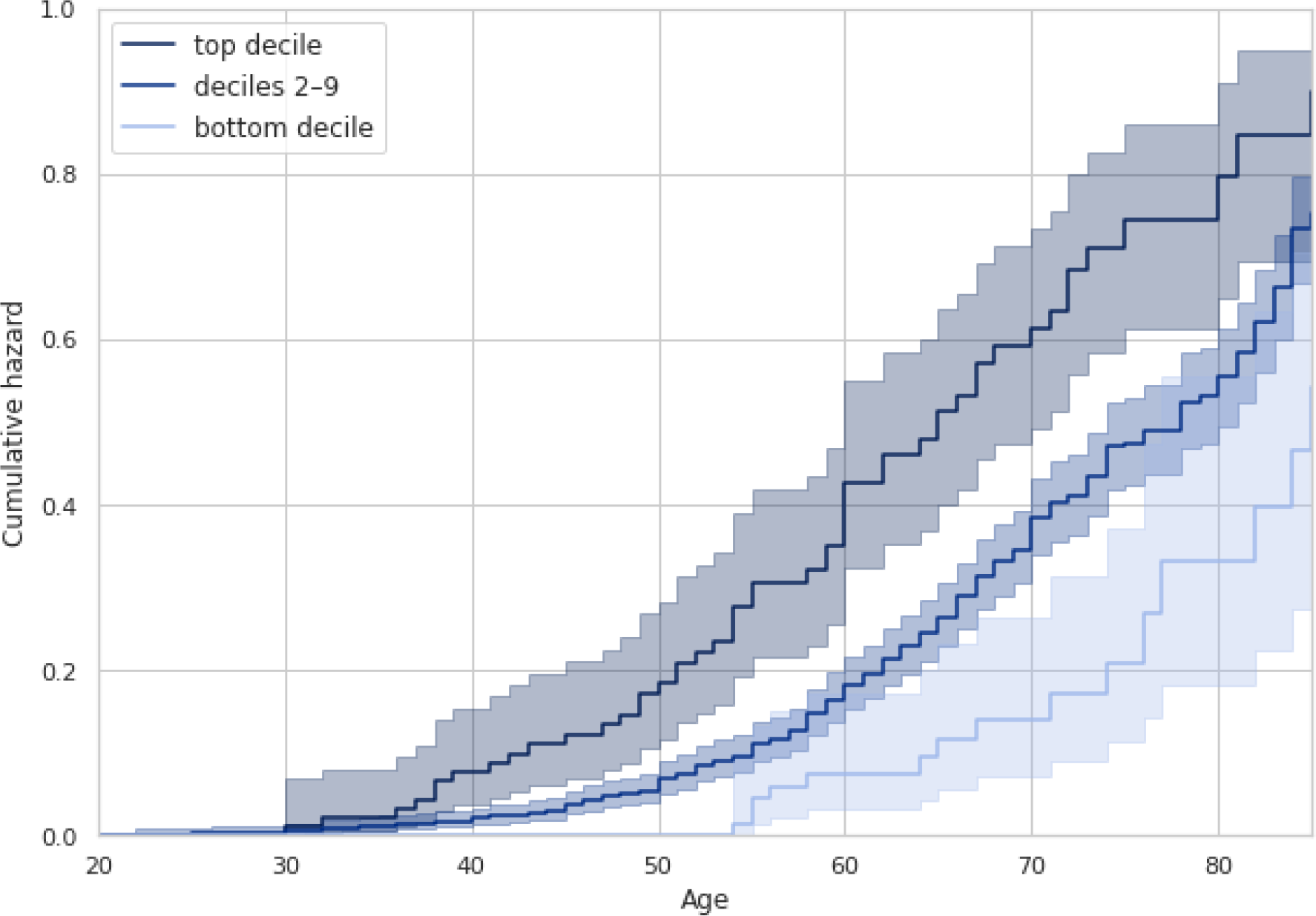
Association of PRS with Incident Severe COVID-19. All participants (N = 1,040) were stratified into three categories, based on their PRS: bottom decile, deciles 2–9, and top decile. Incident severe COVID-19 is plotted according to the PRS category.

### Receiver Operating Curve (ROC) analysis

Next we analysed the association between PRS and severe COVID-19 using a multivariate logistic regression model adjusted for sex, age, and the first 10 principal components of genetic variation. In the adjusted model, a significant association between PRS and severe COVID-19 was found: OR = 1.51 per standard deviation (95% CI: 1.3–1.7 with p-value < 0.0001). High values of PRS (the 10% of PRS distribution) were associated with the adjusted OR = 2.5 (95% CI: 1.6–3.9, p-value < 0.0001, the reference was taken as the mean deciles 10%-90%). In comparison, unadjusted OR = 2.1 (95% CI: 1.4–3.2, p-value = 0.0004 with the reference taken as the mean deciles 10%-90%).

Analyses showed significant (p-value < 0.0001) improvements in AUC with the addition of PRS to the base model containing only the demographic predictors. Figure 6 shows that a model predicting the risk of severe COVID-19 had an AUC of 65% (95% CI: 62–69% by the formula given by Hanley and McNeil [43]) for a model excluding PRS, and it increased up to 67% (95% CI: 64–71%) when PRS was included.

**Figure 6.**
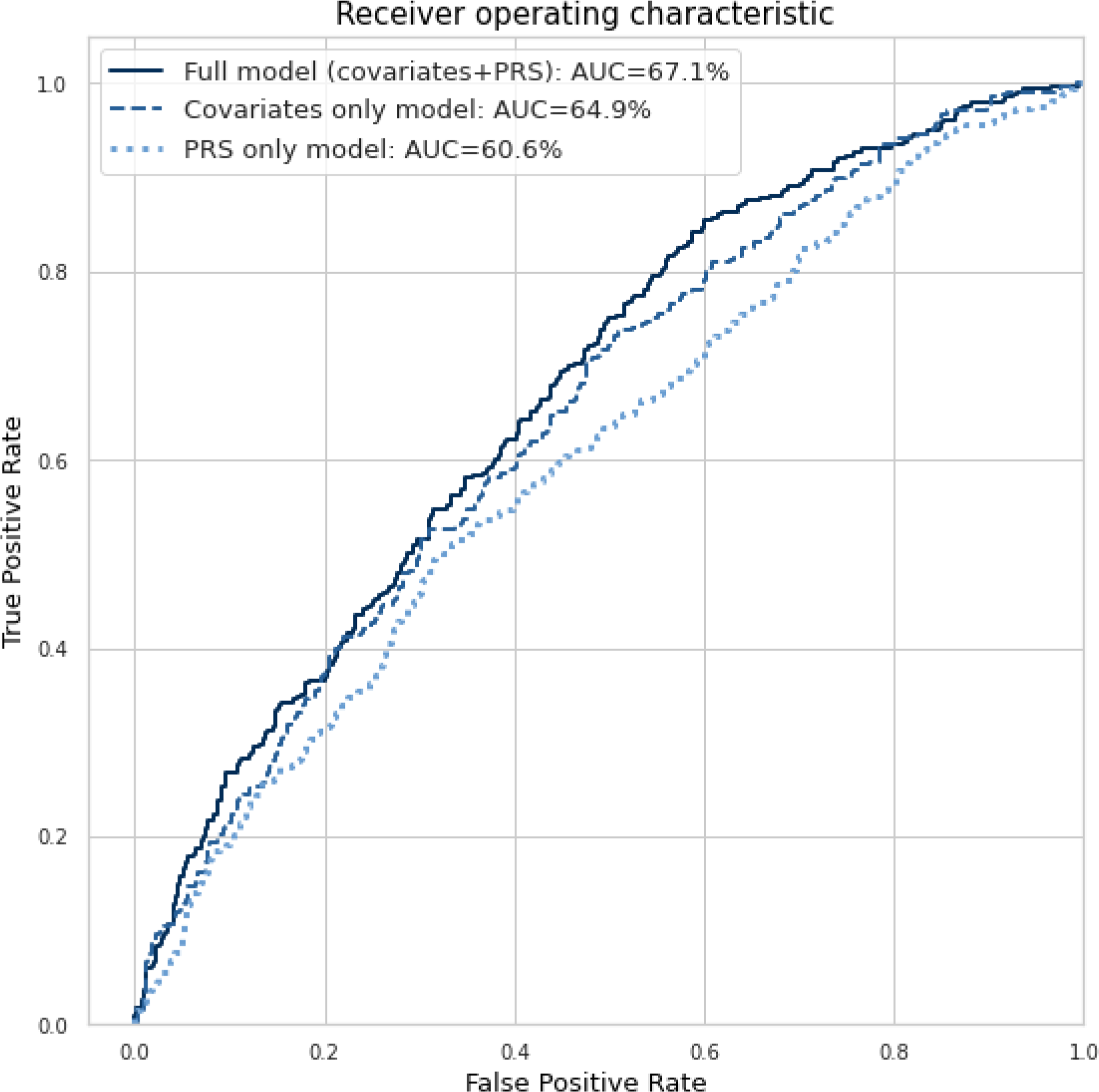
The comparison of receiving operating curves for three logistic regression models. The full model included the demographic predictors (sex and age), the PRS, and the first 10 principal components of genetic variation, while the covariates-only model excluded the PRS.

## DISCUSSION

In this study, we constructed a polygenic risk model for the prediction of the severity of COVID-19 and applied it to a target cohort of 1,040 Russian participants. Comparing the distributions of a PRS, incorporating information from one million common genetic variants between the case and control groups revealed significant differences, indicating meaningful associations between the PRS and corresponding COVID-19 outcomes. We also demonstrated the potential of LP-WGS with coverage less than 5x to be used for predicting the severity of COVID-19.

Our main objective was to evaluate the predictive ability of PRS for COVID-19 severity. To achieve this, we developed a logistic regression model that included only demographic and technical covariates and the full model that also incorporated the PRS. Comparison between these models demonstrated that incorporating our PRS significantly enhanced the predictive accuracy. These findings align with a previous analysis made by Huang et al., where PRS values for severe COVID-19 were constructed by using 112 SNPs in 430,582 participants from the UK Biobank study [29]. In this work, AUC was calculated for a model including only demographic and clinical parameters, and for the full model, which also included the PRS. For the first model, the AUC was 0.789, while in the full mode, the AUC was 0.794 (p-value = 0.002 for increment in AUC). Higher overall prediction accuracy of the model could be attributed to utilisation of information on comorbidities (cardiovascular disease, hypertension, diabetes, chronic respiratory infections, asthma, and chronic obstructive pulmonary disease). Our PRS, based on approximately one million SNPs, gave a comparable improvement in AUC (2% in our study vs 0.5% of Huang et al.). The higher contribution of the PRS in our case can be explained by the much larger number of genetic variants used but also by the absence of clinical factors in our model. Indeed, it is often observed that adding a predictor to a model having a high AUC improves it by an amount smaller than what could be achieved by adding the same factor to a poorer model.

Furthermore, stratifying individuals by PRS quantiles revealed an association with a distinctive risk of severe COVID-19 in resulting groups. The highest PRS categories generally exhibited higher (up to 2.1 for the top 10% PRS) odds ratios. This genetic basis for differences in disease severity among individuals also extended to the occurrence of fatalities due to COVID-19 (OR = 1.8 for the top 10% PRS). These results demonstrate that polygenic risks can be employed to stratify patients and assess their risk of severe disease and mortality related to COVID-19.

Additional survival analysis using the non-parametric Kaplan-Meier estimation revealed that the highest risk categories as defined by PRS not only exhibited higher odds ratios for COVID-19 severity but also experienced an earlier onset of increased risk compared to the mean- and low-risk categories. These findings provide insights into both the overall risk for severe COVID-19 and how the risk varies by age.

These results can have practical implications for protecting individuals with a greater genetic vulnerability during potential future outbreaks. Targeted public health interventions, such as shielding measures, closer monitoring, protection from high-risk frontline work, and prioritization for vaccination, could help to mitigate the associated risk. Hospital-based applications of PRS could facilitate the screening of COVID-19 patients and aid in the early detection of severe disease [28]. Moreover, informing patients about their increased polygenic risk has shown some evidence of positive behavioural impact [44], potentially leading to a decrease in risk-taking behaviours and promoting better outcomes.

A few limitations of the study should be noted. Firstly, despite the multi-ethnic and global nature of the HGI Release 7 meta-analysis, the participants were mostly of Western European descent, which may have affected the accuracy of the predictions in non-Western European populations [9]. Additionally, the lack of detailed clinical data led to the use of CT scans as a criterion for disease severity, which could have introduced some inaccuracy in the classification of the outcome measure for some participants.

## METHODS

### Study population and genetic sequencing

As part of the COVID-19 study, biomaterial (blood) and clinical data from COVID-19 patients hospitalized in the infectious disease department of the St. Petersburg State Budgetary Healthcare Institution “City Hospital No. 40 of Kurortny District” were collected. In this work, low-coverage (2–5x) sequencing was performed for 1040 samples divided into 41 batch sizes. Low-coverage sequencing, also called LP-WGS (low-pass whole genome sequencing), is a low-cost, high-throughput DNA sequencing technology used to accurately detect genetic variation in the genomes of multiple species [45]. Using imputation algorithms, this technology provides high variant detection accuracy with low sequence coverage. LP-WGS and subsequent imputation yield more accurate genotypes than imputation using array-based genotyping data, allowing for increased power in GWAS studies and more accurate results in polygenic risk studies [46].

Prior to sequencing, preliminary analysis and quality control of the case database were performed, and preliminary analysis of samples from each batch was performed to exclude bias for any of the sample characteristics: age, sex, and case/control.

Genome DNA isolation was performed with QIAcube, using a QIAamp DNA Blood Mini Kit. DNA concentration is measured with a Promega QuantiFluor dsDNA System. Library preparation was done using a Roche KAPA HyperPlus Kit. Quality control electrophoresis was done on a QIAxcel station using a QIAxcel High Resolution Kit. Circularization was made with an MGIEasy Circularization Kit. Sequencing was done on a MGISEQ-2000 sequencing machine with the DNBSEQ-G400RS High-throughput Sequencing Set (FCL PE150, 540 G).

### Variant calling, imputation, and quality control

Quality analysis (FastQC, version 0.11.9) [47], alignment (BWA, version 0.7.17) [48], deduplication (samtools, version 1.16.1), and variant collation (bcftools, version 1.16) were performed for the reads obtained from sequencing [49]. Imputation of the resulting data was then performed using the GLIMPSE tool (version 1.1.1) [38], which allows imputation of low-coverage sequencing data. To improve imputation quality, only bi-allelic sites were retained from the LP-WGS BAM data and processed with bcftools. Then iterative refinement of the genotype likelihood using the reference panels with segmentation size of 2 Mb with buffer size of 200 kb produced imputed dosages and multiple chunks within each chromosome were ligated. A panel of the 1000 Genomes Project with high coverage [40], including high-quality single nucleotide variants and insertion-deletion mutations (SNV- and INDELs) from over 3,000 samples, was used as a reference sample.

Subsequently, we filtered imputed variants by an imputation INFO score, where variants with score < 0.7 and a minor allele frequency < 0.1% were removed from the analysis [9,13]. We focused on the variants and individuals with a call rate of more than 90%. Close relatives, identified using the KING-robust method [50], were removed from analysis. Using a threshold (kinship > 0.125), we found pairs of first- and second-degree relatives. We restricted our analyses to a list of variants from HapMap3 [51], which are included in the PRS models. PLINK 1.90 software [52] was utilised for all genotype extraction and quality control.

### Establishing COVID-19 outcomes

The severity of the course was divided according to the following criteria: the case group included samples with lung lesions greater than 50% (computed tomography (CT)-3 and CT-4), while the control group included all other samples. As a result, the case group included 336 patients (208 men and 128 women, 63±15 years) with lung damage more than 50% (computed tomography (CT)-3 and CT-4), the control group included 704 patients (374 men and 330 women, 56±16 years), with lung damage less than 50% or without COVID-19.

### Construction of PRS models

The calculation of PRS values relies on both genotype data from the target individuals and a PRS model. To derive a PRS model, GWAS are used to estimate the effect sizes of SNPs [53]. However, a GWAS gives the marginal effect size for each SNP estimated by a regression model that ignores linkage disequilibrium (LD) structure. As a result, to construct a PRS model that incorporates multiple SNPs, the SNP effects must be re-estimated while accounting for LD structure.

We used the GWAS from the COVID-19 Host Genetics Initiative consortium (release 7). These results were obtained by meta-analysis, which combined the results of 60 individual studies from 25 countries.

To re-weight the effect sizes we used SBayesR (version gctb_2.02), a software tool that has demonstrated superior performance compared to similar tools [19]. This tool re-weights the effects of each genetic variant based on the marginal estimate of its effect size, statistical strength of association, the degree of correlation between the variant and other variants nearby, and tuning parameters. It also requires a GCTB-compatible LD matrix file based on individual-level data from a reference population, and for this analysis, we used a shrunk sparse GCTB LD matrix from 50,000 individuals of European ancestry in the UK Biobank dataset [41].

PRS values were calculated as a weighted sum of allele counts:

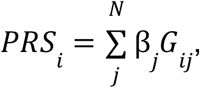

with *βj* the re-weighted effect size of the *j*^*th*^ SNP, *G*_*ij*_ the genotype of the *j*^*th*^ SNP for *i* ^*th*^ individual, *N* the number of SNPs from the model. PLINK 1.90 software [52] was utilised for PRS calculation.

### Statistical analysis and association testing

Logistic regression of PRS categories against COVID-19 severity outcomes was then conducted using R (version 4.3.2) [54] and Python (version 3.8.17) [55], fully adjusted for covariates, such as sex and age. Data on comorbidities were not available for the majority of patients, nor were other clinical data, so parameters for these were not included in the model to cover as much data as possible. The first 10 principal genetic components (PCs) were also included as covariates to adjust for population genetic structures and avoid bias, as per current recommendations [13].

The discriminative power of models in identifying high-risk individuals was then assessed using receiver operating curve (ROC) analysis. The area under the ROC (AUC) was calculated for full models (consisting of covariates and PRS) and base models (covariates only). The confidence interval for the AUC was calculated using the formula given by Hanley and McNeil [43]. Increment in AUC (ΔAUC) was reported based on the difference between the two models, reported as the discriminative or predictive power conferred by PRS. The permutation test for differences between classifiers was used to estimate the significance (p-value) of an increment in AUC.

Once the PRS was calculated, individuals were separately stratified into quintiles for susceptibility and severity PRS, then categorised into low genetic risk (decile 1, bottom 10% of cohort), intermediate risk (decile 2–9, middle 80%) and high risk (decile 10, top 10%) for each outcome. In each group, we estimated the cumulative hazard curve using the non-parametric method called the Kaplan-Meier estimator [56]. For each pair of groups, the log rank test was applied, which is the statistical test for comparing the survival distributions of two or more groups.

## Data Availability

All data produced in the present study are available upon reasonable request to the authors.

## DATA AND CODE AVAILABILITY

Personal genetic and clinical data are under restrictions and are available through collaboration with the St. Petersburg State Health Care Institution “City Hospital No. 40, Kurortny District” hospital.

### ACKNOWLEDGEMENTS

The authors are grateful to the study participants and the staff from the St. Petersburg State Health Care Institution “City Hospital No. 40, Kurortny District” hospital. The authors would like to thank all authors of the included studies for their valuable contributions to data collection. This work was supported by Saint Petersburg State University, project ID: 94029859.

## ETHICS STATEMENT

Ethics committee of The Saint Petersburg State Health Care Establishment the City Hospital No 40 of the Resort District gave ethical approval for this work (protocol number 171, May 18, 2020).

