## supplementary_material for "Analysis of associations between polygenic risk score and COVID-19 severity in a Russian population using low-pass genome sequencing"

**Supplementary Materials**

These supplementary materials present additional details about the results.

### LOGISTIC REGRESSION RESULTS

#### The full model

We analysed the association between PRS and severe COVID-19 using a multivariate logistic regression model adjusted for sex, age, and the first 10 principal components of genetic variation.

**Table 1:** Logistic regression results for the full model.

| | coef | std err | z | P>\|z\| | [0.025 | 0.975] |
|---|---|---|---|---|---|---|
| **intercept** | -0.0495 | 0.219 | -0.226 | 0.821 | -0.478 | 0.379 |
| **age** | 0.5637 | 0.078 | 7.187 | 0.000 | 0.410 | 0.717 |
| **sex** | -0.5737 | 0.145 | -3.952 | 0.000 | -0.858 | -0.289 |
| **PRS** | 0.4135 | 0.073 | 5.651 | 0.000 | 0.270 | 0.557 |
| **PC1** | -0.0025 | 0.095 | -0.026 | 0.979 | -0.189 | 0.184 |
| **PC2** | 0.2154 | 1.240 | 0.174 | 0.862 | -2.216 | 2.647 |
| **PC3** | -0.5792 | 1.203 | -0.481 | 0.630 | -2.938 | 1.779 |
| **PC4** | 0.3239 | 0.599 | 0.541 | 0.589 | -0.850 | 1.498 |
| **PC5** | 0.3810 | 0.409 | 0.931 | 0.352 | -0.421 | 1.183 |
| **PC6** | 0.2982 | 0.296 | 1.009 | 0.313 | -0.281 | 0.878 |
| **PC7** | -0.1118 | 0.109 | -1.022 | 0.307 | -0.326 | 0.103 |
| **PC8** | -0.0367 | 0.072 | -0.510 | 0.610 | -0.178 | 0.104 |
| **PC9** | 0.0875 | 0.085 | 1.031 | 0.303 | -0.079 | 0.254 |
| **PC10** | 0.0153 | 0.072 | 0.212 | 0.832 | -0.127 | 0.157 |

- **coef**: the coefficient of the variable (log[odd ratio]);
- **std err**: the standard error of the coefficient;
- **Z**: the z value for the estimated coefficient and the standard error;
- **P>|z|**: the p-value associated with the value in the z value column;
- **[0.025, 0.975]**: 95% confident interval (CI) for the coefficient.

#### The covariates-only model

The base model contains only the demographic predictors (sex, age, and the first 10 principal components of genetic variation).

**Table 2:** Logistic regression results for the covariates-only model.

| | coef | std err | z | P>\|z\| | [0.025 | 0.975] |
|---|---|---|---|---|---|---|

| | | | | | | |
|---|---|---|---|---|---|---|
| **intercept** | -0.0549 | 0.214 | -0.257 | 0.797 | -0.474 | 0.364 |
| **age** | 0.5321 | 0.076 | 6.982 | 0.000 | 0.383 | 0.681 |
| **sex** | -0.5383 | 0.143 | -3.775 | 0.000 | -0.818 | -0.259 |
| **PC1** | -0.0284 | 0.093 | -0.305 | 0.760 | -0.211 | 0.154 |
| **PC2** | 0.8426 | 1.226 | 0.687 | 0.492 | -1.561 | 3.246 |
| **PC3** | -1.0866 | 1.186 | -0.916 | 0.360 | -3.412 | 1.238 |
| **PC4** | 0.2649 | 0.513 | 0.516 | 0.606 | -0.741 | 1.271 |
| **PC5** | 0.3364 | 0.354 | 0.950 | 0.342 | -0.357 | 1.030 |
| **PC6** | 0.2184 | 0.250 | 0.874 | 0.382 | -0.271 | 0.708 |
| **PC7** | -0.0971 | 0.117 | -0.832 | 0.405 | -0.326 | 0.132 |
| **PC8** | -0.0362 | 0.071 | -0.508 | 0.612 | -0.176 | 0.104 |
| **PC9** | 0.1146 | 0.083 | 1.377 | 0.169 | -0.049 | 0.278 |
| **PC10** | 0.0180 | 0.071 | 0.255 | 0.799 | -0.121 | 0.157 |

**The PRS-only model**

The base model contains only PRS predictor.

**Table 3:** Logistic regression results for the PRS-only model.

| | **coef** | **std err** | **z** | **P>\|z\|** | **[0.025** | **0.975]** |
|---|---|---|---|---|---|---|
| **intercept** | -0.8025 | 0.081 | -9.923 | 0.000 | -0.961 | -0.644 |
| **PRS** | 0.3727 | 0.071 | 5.278 | 0.000 | 0.234 | 0.511 |
| **PC1** | -0.0165 | 0.092 | -0.178 | 0.858 | -0.197 | 0.164 |
| **PC2** | 0.4333 | 1.177 | 0.368 | 0.713 | -1.874 | 2.740 |
| **PC3** | -0.8196 | 1.152 | -0.711 | 0.477 | -3.078 | 1.439 |
| **PC4** | 0.2147 | 0.525 | 0.409 | 0.683 | -0.815 | 1.244 |
| **PC5** | 0.3026 | 0.358 | 0.845 | 0.398 | -0.399 | 1.005 |
| **PC6** | 0.2557 | 0.263 | 0.972 | 0.331 | -0.260 | 0.771 |
| **PC7** | -0.1390 | 0.111 | -1.254 | 0.210 | -0.356 | 0.078 |
| **PC8** | -0.0669 | 0.077 | -0.869 | 0.385 | -0.218 | 0.084 |
| **PC9** | 0.0808 | 0.083 | 0.977 | 0.328 | -0.081 | 0.243 |
| **PC10** | -0.0271 | 0.070 | -0.387 | 0.699 | -0.164 | 0.110 |

**The full model with PRS as a binary predictor**

We analysed the association between PRS and severe COVID-19 using a multivariate logistic regression model adjusted for sex, age, and the first 10 principal components of genetic variation. PRS values were considered as a binary predictor, where the highest 10% of PRS were taken as 1, others as 0.

**Table 4:** Logistic regression results for the full model with PRS as a binary predictor.

| | coef | std err | z | P>\|z\| | [0.025 | 0.975] |
|---|---|---|---|---|---|---|
| **intercept** | -0.0781 | 0.223 | -0.350 | 0.726 | -0.515 | 0.359 |
| **10% with the highest PRS values** | 0.9143 | 0.225 | 4.069 | 0.000 | 0.474 | 1.355 |
| **10% with the lowest PRS values** | -1.0727 | 0.290 | -3.703 | 0.000 | -1.640 | -0.505 |
| **age** | 0.5789 | 0.079 | 7.342 | 0.000 | 0.424 | 0.733 |
| **sex** | -0.5430 | 0.145 | -3.746 | 0.000 | -0.827 | -0.259 |
| **PC1** | -0.0078 | 0.095 | -0.082 | 0.935 | -0.195 | 0.179 |
| **PC2** | 0.5737 | 1.249 | 0.459 | 0.646 | -1.875 | 3.023 |
| **PC3** | -0.8886 | 1.208 | -0.736 | 0.462 | -3.256 | 1.479 |
| **PC4** | 0.3058 | 0.612 | 0.500 | 0.617 | -0.893 | 1.505 |
| **PC5** | 0.3584 | 0.417 | 0.859 | 0.390 | -0.459 | 1.176 |
| **PC6** | 0.3072 | 0.300 | 1.023 | 0.306 | -0.281 | 0.896 |
| **PC7** | -0.0974 | 0.112 | -0.871 | 0.384 | -0.317 | 0.122 |
| **PC8** | -0.0315 | 0.072 | -0.436 | 0.663 | -0.173 | 0.110 |
| **PC9** | 0.0818 | 0.085 | 0.959 | 0.338 | -0.085 | 0.249 |
| **PC10** | 0.0190 | 0.073 | 0.261 | 0.794 | -0.123 | 0.161 |
